# Selecting the most effective DBS contact in essential tremor patients based on individual tractography

**DOI:** 10.1101/2020.11.03.20211847

**Authors:** Jan Niklas Petry-Schmelzer, Till A. Dembek, Julia K. Steffen, Hannah Jergas, Haidar S. Dafsari, Gereon R. Fink, Veerle Visser-Vandewalle, Michael T. Barbe

## Abstract

Postoperative choice of the most effective DBS contact in patients with essential tremor (ET) so far relies on lengthy clinical testing. It has been shown that the postoperative effectiveness of DBS contacts depends on the distance to the dentatorubrothalamic tract (DRTT). Here, we investigated whether the most effective DBS contact could be determined from the stimulation overlap with the individual DRTT. Seven ET patients with bilateral thalamic deep brain stimulation were included retrospectively. Tremor control was assessed contact-wise during test stimulation with 2mA. The individual DRTTs were identified from diffusion tensor imaging. Contacts were ranked by their overlap of the test stimulation with the respective DRTT in relation to their clinical effectiveness. A linear mixed-effects model was calculated to determine the influence of the DRTT-overlap on tremor control. In 92.9 % of the cases, the contact with the best clinical effect was the contact with the highest or second-highest DRTT-overlap. On the group level, the DRTT-overlap explained 26.7% of the variance of the clinical outcome (p<0.001). To conclude, data suggest that the overlap with the DRTT based on individual tractography may serve as a marker to determine the most effective DBS contact in ET patients and reduce burdensome clinical testing in the future.

## 1. Introduction

Essential tremor (ET) is the most common adult movement disorder, causes significant disability, interferes with activities of daily living, and reduces quality of life. For medication-refractory cases, deep brain stimulation (DBS) of the thalamic ventral intermediate nucleus (VIM) and the posterior subthalamic area (PSA) is an established effective, and safe treatment [1,2]. However, postoperative choice of the most effective contact relies on time-consuming and exhausting clinical testing, especially with new generations of “directional leads” consisting of up to eight contacts.

DBS most likely modulates pathologic activity within the tremor network via cerebello-thalamo-cortical connections, i.e., the dentatorubrothalamic tract (DRTT) [3]. Additionally, it has been shown that direct targeting of the DRTT leads to successful tremor control and that the effectiveness of a contact depends on its distance to the DRTT [3–6]. We hypothesized that the most effective postoperative contact can be determined in silico by the overlap of the stimulation with the respective DRTT.

## 2. Materials and Methods

### 2.1. Study design

This retrospective study included ET patients with bilateral stereotactic DBS lead implantation in PSA/VIM, who had undergone neurosurgery between January 2019 and January 2020, with available preoperative cerebral MRI, diffusion tensor imaging (DTI), and postoperative CT. Patient selection and implantation procedures have been described in detail [2]. All patients were implanted with directional leads (Cartesia^™^, Boston Scientific, USA). Three months postoperatively, patients underwent routine clinical testing to determine the most effective contact for postoperative tremor control. The study was approved by the local ethics committee (Vote: 20-1511). Due to the retrospective character of the study no informed consent was needed.

### 2.2. Clinical outcome and lead reconstruction

As per clinical routine at our center, postural tremor, intention tremor, and rest tremor of the upper limb contralateral to the active stimulation site were assessed without stimulation and during contact-wise stimulation with a fixed amplitude of 2mA, a frequency of 130Hz, a pulse width of 60µs. Contralateral stimulation was switched off during the testing. Clinical scores ranged from 0 (“no tremor”) to 4 (“most severe tremor”). For directional levels, each contact was examined separately. For further analysis, the sum of postural, intention, and rest tremor percentual change scores from the “OFF stimulation” baseline were calculated. DBS leads and their respective rotation were identified from postoperative CT scans, and lead locations were transformed into the preoperative MRI using the Lead-DBS toolbox (www.lead-dbs.org) [7–9]. Respective volumes of tissue activated (VTAs) were calculated in individual patient space using FASTFIELD with an electrical field threshold of 0.2 V/mm and an isotropic conductivity of 0.1 S/m [10,11].

### 2.3. Probabilistic tracking of the DRTT

MRI data were acquired on a 3-Tesla Philips Ingenia® Scanner (Philips, Amsterdam, The Netherlands) (T1-sequence: TR: 9.8ms, TE: 4.9ms, acquisition time: 6:13min, voxel-size: 0.49×0.49×1.00mm^3^). For diffusion imaging, a single-shot 2D, spin-echo, echo-planar imaging pulse sequence was applied (TR: 8213ms, TE: 103ms, 40 gradient directions, b-value: 1000s/mm^2^, acquisition time: 9:53min, voxel-size; 2.0×2.0×2.0mm^3^). For probabilistic fiber tracking, we used the FMRIB software library (FMRIB, Oxford, UK). We employed probabilistic fiber tracking, as it might be better in detecting the DRTT than the deterministic algorithms embedded in commercially available stereotactic planning software [12]. Diffusion data were corrected for susceptibility-induced distortions using the topup-tool and corrected for head motion and eddy current distortion using the eddy-tool. Brain extraction of the b0-image was performed using the BET-tool, and distributions of diffusion vectors were estimated for each voxel with BEDPOSTX. The number of fibers per voxel was set to two. Probabilistic fiber tracking was performed separately for each DRTT with PROBTRAACKX2 using modified Euler integration. For all other parameters, the respective default settings were used. The choice of regions of interest has been described previously [5]. In brief, the contralateral dentate nucleus was chosen as a seed region, while the contralateral superior cerebellar peduncle, the ipsilateral red nucleus, and the ipsilateral precentral gyrus served as waypoints. These regions of interest were previously defined in MNI space and transformed to individual diffusion space using SPM (http://www.fil.ion.ucl.ac.uk/spm/software/spm12/) [5]. The resulting track frequency maps were visually examined for anatomical accuracy and transformed into a track probability map, according to Schlaier et al. [12]. Finally, the resulting fiber tracts were coregistered to the preoperative T1.

### 2.4. Statistical analysis

For further analysis, the overlap of each 2mA VTA with the respective DRTT was calculated as the sum of the resulting track probability map values covered by the respective VTA, multiplied with the VTA’s voxel-size in mm^3^. These DRTT-overlap values were ranked hemisphere-wise to determine the contact with the largest DRTT-overlap. We then investigated how often the electrodes with the highest and second-highest overlap also had the best clinical outcome during clinical testing. Additionally, a linear mixed-effects model was employed to determine the predictive value of the overlap with the individual DRTT regarding tremor control on the group level. We included “DRTT overlap” as the main effect and “lead” as a random-effect, to take multiple testing per lead into account. [13].

### 2.5. Data availability

Matlab scripts are available from the open science framework (Link follows on acceptance). Anonymized imaging and clinical data are available upon request to the corresponding author and not publicly available due to privacy concerns.

## 3. Results

A total of 7 ET patients (3 female, age: 68.8 y ±14.8) and 14 directional DBS leads were included in this retrospective analysis. The contact ranking results by clinical effectiveness and overlap with the individual DRTT are shown in Figure 1. In 71.4 % of the cases (10 of 14 hemispheres), the contact with the highest overlap with the individual DRTT showed the best clinical outcome or was among those with the best outcome if more than one contact showed equal tremor improvement. In the remaining cases, in 3 of 4 hemispheres the contact with the second-highest overlap with the individual DRTT showed the best clinical outcome. When only investigating directional contacts, in 64.3 % of the cases the directional contact with the highest DRTT-overlap also had the best tremor improvement. On the group level, the linear mixed-effects model explained 68.4% of the variance of clinical outcome, while the overlap with the individual DRTT alone (main-effect) explained 26.7% of the variance (R^2^_model_ = 0.684, R^2^_main-effect_ = 0.267, p<0.001, see Figure 2). A positive relationship between DRTT-overlap and tremor improvement was observed in all hemispheres.

**Figure 1.**
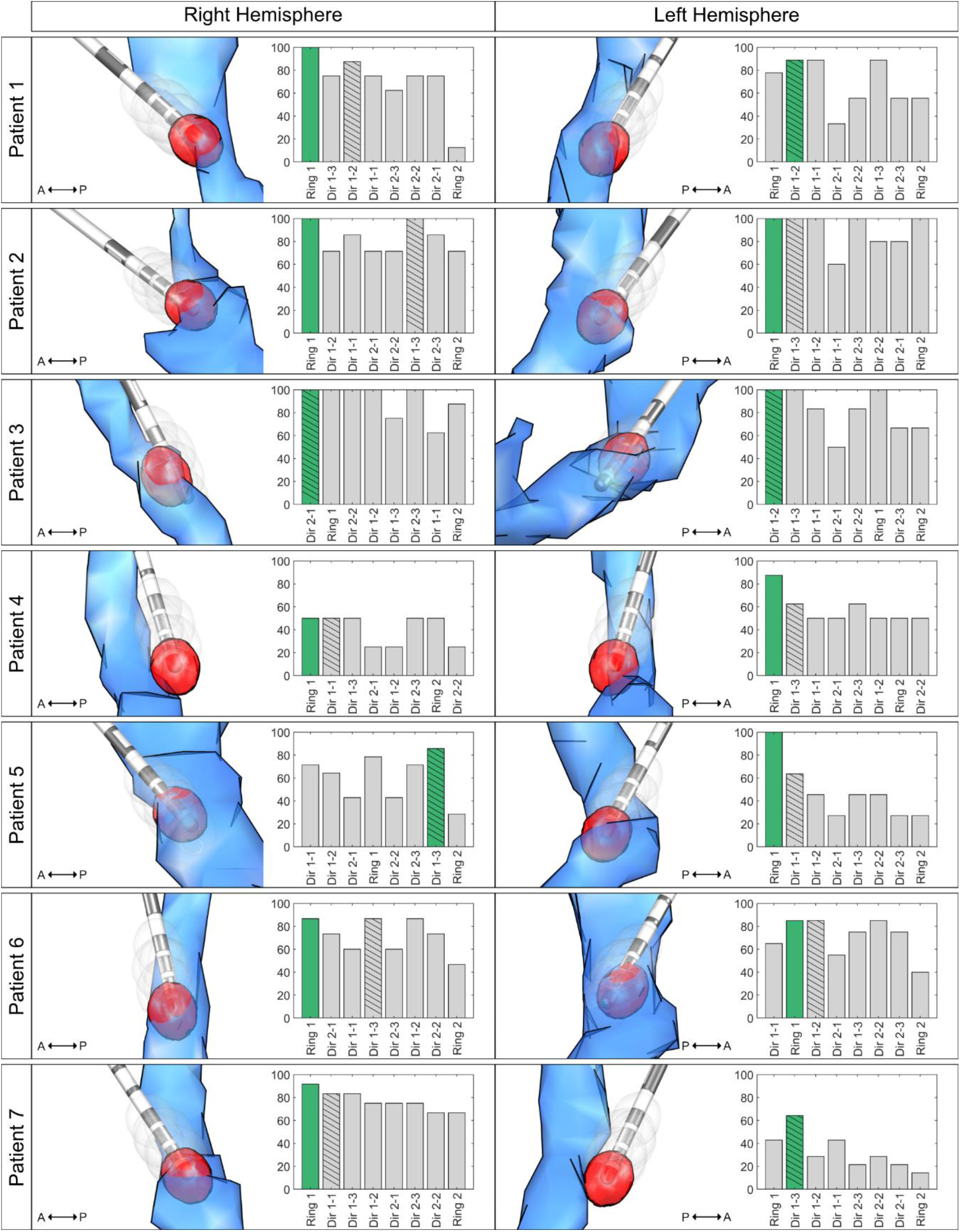
Contact Ranking. Bar plots illustrate the ranking of the overlap with the individual dentatorubrothalamic tract (DRTT) per contact on the x-axis (highest overlap to lowest overlap) and the improvement in tremor control in % on the y-axis. The most effective contact is marked in green (with the highest overlap in case more than one contact had the best improvement), and the bar of the most effective directional contact is hatched. The respective left column illustrates the relation of the generated volumes of tissue activated (VTAs, gray) together with the DRTT (blue) and the respective lead in the medial view. The VTA with the highest DRTT-overlap is highlighted in red. For illustration, only the 10% highest values of the track probability map are shown. **Abbreviations:** A = anterior, DRTT = dentatorubrothalamic tract, P = posterior, VTA = volume of tissue activated, Dir 1 = ventral directional level, Dir 2 = dorsal directional level.

**Figure 2.**
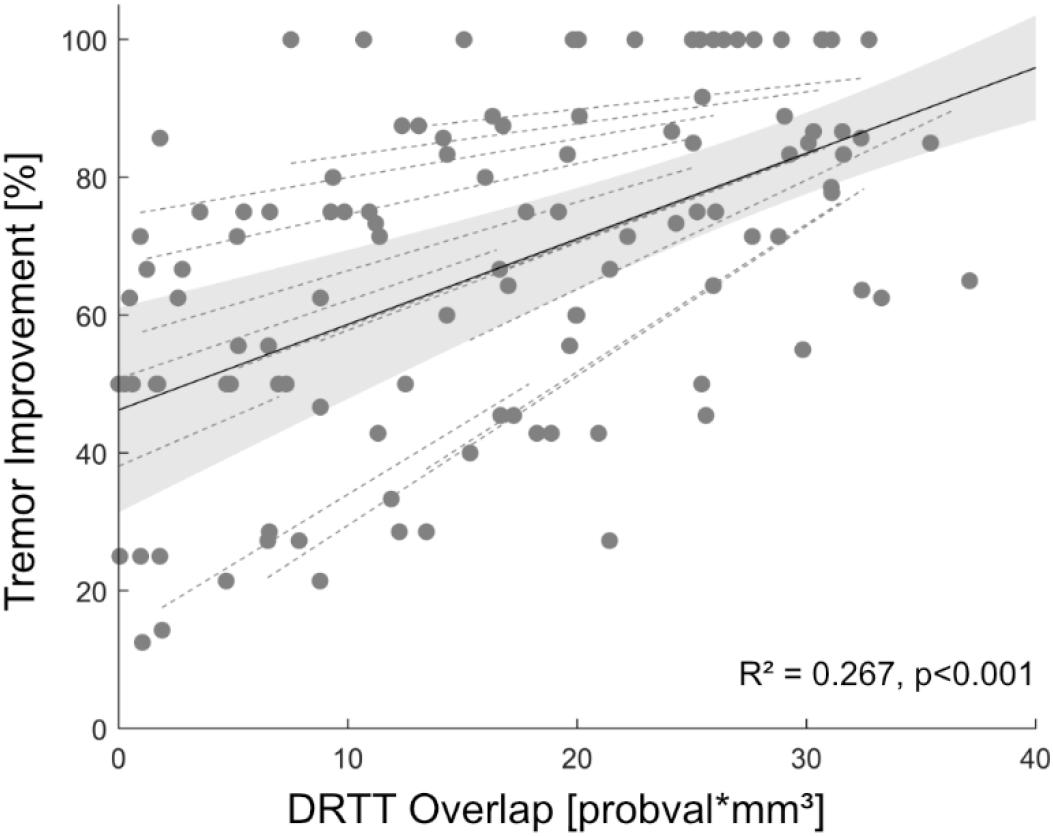
Prediction of Tremor Improvement. Linear mixed-effects model (black) and 95% confidence interval (gray) between tremor improvement and overlap with the individual dentatorubrothalamic tract (DRTT). Random effects for each individual hemisphere are also shown (dashed, gray). **Abbreviations:** DRTT=dentatorubrothalamic tract.

## 4. Discussion

This study demonstrates that the overlap with the DRTT might serve as a marker for in silico determination of the most effective contact for tremor suppression in ET patients. The overlap with the DRTT determined one of the most effective contacts in 71.4%. When also considering the contact with the second-most overlap, this figure increased to 92.9% of the cases. In other words, if one had only interrogated the two contacts with the highest DRTT-overlap, a contact with an optimal outcome would have been determined in 13/14 hemispheres. In the remaining hemisphere (Patient 5, right hemisphere) in which the most effective contact ranked worse, i.e., in seventh place, the contact with the highest overlap still was on the same directional level as the most effective contact and improved tremor by 75%. With new generations of DBS leads, there is the option of steering the current towards more effective contacts, away from contacts causing side effects [14]. Only considering directional contacts, the chance of activating the most effective directional contact without clinical testing increases from 16.7% (1 out of 6) to 64.3% when using the in silico approach presented here.

Although landmark-based targeting was used when implanting our patients [2], in the majority of cases, the most ventral contact was the most effective contact with the highest overlap to the DRTT. This finding is in line with previous studies indicating that (i) effective contacts are located inside or close to the DRTT and [3,5] (ii) that in our targeting approach, contacts in the PSA are closer to the DRTT [5]. While there was a positive relationship between DRTT-overlap and tremor suppression in all hemispheres in our mixed-effects model, and overlap predicted 26.7% of the variance in tremor outcome, there were still marked individual differences between hemispheres, as indicated by the 68% of variance explained when also considering hemisphere as a random effect.

Several attempts to predict postoperative tremor suppression focused on connectivity analysis or probabilistic stimulation mapping [15–17]. However, only Åström et al. [18] focused on predicting the DBS contact to be chosen postoperatively. Based on probabilistic stimulation maps, the resulting software tool, showed that the predicted contact with rank 1 matched the clinically used contact in 60% of cases (rank 1-2 matched 83% of the cases). In contrast, the present study was based on the individual DRTT as a neuroanatomical correlate of tremor suppression and included leads with eight contacts instead of four contacts.

This study’s major limitation is that we only investigated tremor suppression and did not consider stimulation-induced side effects. In several cases, more than one contact showed equally optimal tremor suppression. In such cases, side effect thresholds would be crucial for determining the contact used for clinical stimulation. Therefore, more research regarding the neuroanatomical origins of different stimulation-induced side effects [19] is needed. Prospective studies, also taking stimulation-induced side effects such as muscle contractions, paresthesia, ataxia, and stimulation-induced dysarthria into account, should be conducted to validate and extend this retrospective analysis.

Nevertheless, our study demonstrates how in silico imaging analysis could guide clinical DBS programming in ET and help reduce patient burden by shortening tedious monopolar review investigations.

## Data Availability

Data will be available upon acceptance of the manuscript via the openscience framework.

## Supplementary Materials

None.

## Author Contributions

Conceptualization, JNPS and TAD.; methodology, JNPS and TAD; software, JNPS and TAD.; validation, JNPS and TAD; formal analysis,JNPS and TAD; investigation, JNPS and TAD; resources, JKS, HJ, and VVV; data curation, JNPS and TAD; writing—original draft preparation, JNPS and TAD.; writing— review and editing, JKS, HJ, HSD, GRF, VVV, and MTB; visualization, JNPS and TAD.; supervision, HSD, VVV and MTB; project administration, GRF, VVV and MTB.. All authors have read and agreed to the published version of the manuscript.

## Acknowledgments

None

### Appendix A

None.

### Appendix B

None.

